# Trends and Disparities in Newer GLP1 Receptor Agonist Initiation among Real-World Adult Patients Eligible for Obesity Treatment

**DOI:** 10.1101/2025.01.20.25320839

**Authors:** Rotana M. Radwan, Yao An Lee, Pareeta Kotecha, Davene R. Wright, Inmaculada Hernandez, Ronald Ramon, William T. Donahoo, Yong Chen, John M. Allen, Jiang Bian, Jingchuan Guo

**Author notes:** Corresponding Author: Jingchuan Guo, MD Assistant Professor, Department of Pharmaceutical Outcomes and Policy University of Florida College of Pharmacy, 1889 Museum Road, DSIT 6004, Gainesville, FL, 32606. Conflict of Interest: **None.**.

## Abstract

**Aims:** To characterize trends in the initiation of newer anti-obesity medications (AOMs) and determine factors associated with their use among obese/overweight populations.

**Materials and methods:** This retrospective study utilized electronic health record data from OneFlorida+ (2015–2024). Adults eligible for AOMs were included, defined as having a BMI ≥30 kg/m² or a BMI of 27–29.9 kg/m² with at least one obesity-related comorbidity. The primary outcome was the initiation of newer AOMs, specifically glucagon-like peptide-1 receptor agonists (GLP-1 RAs) including liraglutide, semaglutide, and tirzepatide. Trends across years were examined, and a multivariable logistic regression identified sociodemographic, clinical, and healthcare utilization factors associated with AOM initiation.

**Results:** Of 319,949 adults, 1.8% initiated newer AOMs. Semaglutide accounted for 77.9% of initiations, tirzepatide 19.7%, and liraglutide 17.8%. Initiation trends showed liraglutide uptake peaked at 5% in 2018 but declined afterward, while semaglutide and tirzepatide uptake increased exponentially since 2022. Odds of initiation were lower for Black (aOR (95% CI): 0.87 [0.80– 0.94]) and Hispanic (0.84 [0.78–0.91]) groups vs. Whites, and for Medicaid (0.69 [0.63–0.76]) and uninsured (0.81 [0.74–0.87]) patients vs. privately insured. Higher odds were associated with being female, middle-aged, having more outpatient visits, and visiting endocrinologists.

**Conclusions:** The initiation of newer AOMs among overweight and obese populations remains low, but uptake has increased exponentially since 2022. Our findings reveal significant disparities in obesity care, highlighting the importance of addressing inequities in AOM access to improve obesity outcomes.

## Background

Obesity is a multifactorial condition resulting from a complex interaction of environmental, genetic, and behavioral factors (1, 2). Obesity contributes to the global burden of disease and disability by increasing the risk of heart disease, diabetes, hypertension, osteoarthritis, and certain cancers, among other conditions (1). In the United States (US), 41.9% of adults have obesity, and projections show that one in every two adults will have obesity by 2030 (3).

According to clinical guidelines, individuals with a body mass index (BMI) ≥30 kg/m² or BMI ≥27 kg/m² with at least one obesity-associated comorbidity can be offered pharmacotherapy in addition to lifestyle interventions to help achieve their weight-loss goals (4, 5). Historically, anti-obesity medications (AOM) such as orlistat (ALLI/Xenical), phentermine/topiramate (Qsymia), bupropion/naltrexone (Contrave), and lorcaserin (Belviq) have demonstrated an average body weight loss of ≤10% after one year, but with serious safety concerns such as cardiotoxicity, neuropsychiatric disorders, and cancer (6). Recently approved by the US Food and Drug Administration (FDA), glucagon-like peptide-1 receptor agonists (GLP-1 RA), including liraglutide (Saxenda), semaglutide (Wegovy), and tirzepatide (Zepbound), have shown greater effectiveness, health benefits, and safety profiles (7; 8). For example, semaglutide demonstrated up to 15% mean weight loss (8), sustained over at least two years (9), and tirzepatide, showed up to 22% mean weight loss after 72 weeks, along with improved cardiometabolic risk factors, such as blood pressure, insulin levels, and lipid levels (10).

Despite the high prevalence of obesity, the use of AOMs remains relatively low in the US. Between 2015 to 2023, only 8.0% of adults had AOM prescriptions, and just 4.4% filled their prescriptions. Among those who filled prescriptions, 39% received naltrexone/bupropion, 29% phentermine-topiramate, 19% semaglutide, 11% liraglutide, and 1.2% orlistat (11). The initiation of AOMs in clinical practice requires balancing the benefits of treatment with the risks of adverse effects and costs (12). The disproportionate burden of obesity on underserved populations, combined with the lack of coverage under public health plans, raise significant concerns about exacerbating health disparities in AOM access (12). Findings from Gasoyan et al., using data from 2015 to 2023, underscore these concerns by revealing significant disparities in AOM access at both the prescribing and prescription-filling stages, influenced by patient characteristics and insurance type (11). Specifically, non-Hispanic Whites had higher odds of being prescribed an AOM compared to non-Hispanic Blacks, Hispanics, and other racial or ethnic groups, and individuals with private insurance were more likely to receive both AOM prescriptions and have them filled (11).

Limited data exists on the initiation of newer AOMs (e.g., liraglutide, semaglutide, and tirzepatide). Understanding which patient populations are more likely to initiate these treatments may improve obesity management by prioritizing high-risk groups (12). This knowledge also informs health policies regarding insurance coverage and access to obesity specialists, which are vital for population-level obesity management (12). Thus, this study aimed to assess trends in the initiation of newer AOMs and to characterize the patient populations starting these treatments among overweight and obese adults in the US.

## Materials and methods

### Study design, database, and sample

This retrospective cohort study utilized data from the OneFlorida+ Data Trust, a centralized patient data repository that includes longitudinal electronic health records (EHRs) (13). The study included adults aged 18 years or older who were eligible for AOMs, defined as a BMI of 30 kg/m² or higher, or a BMI between 27 and 29.9 kg/m² with at least one weight-related comorbidity between January 1, 2015, and January 31, 2024. The cohort entry date (index date) was the first date a patient was identified as being eligible for AOMs within the study period.

The baseline period was the 12 months preceding the index date, and the follow-up period extended for 12 months after the index date. We included patients who had at least one encounter during the baseline period and follow-up period, respectively. Patients were excluded if they were pregnant, had an active malignant neoplasm, or had used any AOM during the baseline period. This study was approved as exempt by the University of Florida Institutional Review Board.

### Study Outcome

The primary outcome was the initiation of newer AOMs (yes or no) during the follow-up year, including liraglutide, semaglutide, and tirzepatide. The secondary outcome was the initiation of any AOMs, including both newer and traditional AOMs (i.e., orlistat, phentermine/topiramate, bupropion/naltrexone, and lorcaserin).

### Covariates

Baseline covariates were collected within the 1-year period prior to the index date. Demographic and socioeconomic factors included: age (18-40, 40-65, and ≥65), sex (female, male), race or ethnicity (Non-Hispanic White, Non-Hispanic Black, Hispanic, and other), insurance coverage (Medicare, Medicaid, private insurance, uninsured, and other), and area deprivation index (ADI). The ADI is a validated measure of neighborhood adversity based on 17 variables related to education, employment, housing, and income from US Census and American Community Survey data (14, 15). The ADI reflects the average socioeconomic characteristics of a population within a neighborhood. State-level ADIs are available in deciles from 1 to 10, ranking from least disadvantaged (1) to most disadvantaged (10) (14, 15). For this study, ADI was grouped into three decile categories: 1-2 (low ADI), 3-8 (median ADI), and 9-10 (high ADI).

Clinical factors included obesity status (overweight [BMI 27-29 kg/m^2^] and obese [BMI >=30 kg/m^2^]), smoking status (current, previous, and never smoker), and Charlson comorbidity index score (0, 1, and 2+) (16). Comorbidities and medication use were coded as indicator variables for the following: coronary artery disease, heart failure, type 2 diabetes (T2D), arthritis, chronic obstructive pulmonary disease, anxiety, depression, antihypertensives, statins, metformin, other glucose-lowering drugs, opioids, and antidepressants except for bupropion. Additional factors included provider specialty (cardiology, endocrinology, primary care, and specialized nurse practitioner/physician assistant), and healthcare utilization in the baseline year, including the number of outpatient visits (0, 1-2, and 3+), hospitalizations (0, 1+), and emergency room visits (0, 1+).

### Statistical analysis

Sample characteristics were described using categorical data, presented as frequencies and proportions for the overall eligible cohort and subgroups, including newer AOM initiators, traditional AOM initiators, and non-AOM initiators. The proportion of initiating newer AOM for each drug was calculated by calendar year, and trends were plotted. Additionally, the incidence proportion of initiating newer AOMs, both overall and by individual drugs, was calculated across five-digit ZIP code regions in Florida. Geographic variation in newer AOM initiation was visualized through maps covering the entire study period (January 1, 2015, to January 31, 2024) and specifically highlighting the years 2022 and 2023.

To identify factors associated with the initiation of newer AOMs (yes/no) or any AOMs (yes/no) among the eligible cohort, two multivariable logistic regressions were conducted. The dependent variables were the initiation of newer AOMs and the initiation of any AOMs, respectively.

Independent variables included age, sex, race and ethnicity, insurance coverage, ADI, comorbidities, medication use, provider specialty, and healthcare utilization data, such as outpatient visits, emergency visits, and hospitalizations.

Subgroup analyses were conducted to identify factors associated with the initiation of newer AOMs among individuals with and without T2D, as well as among those who initiated treatment in 2023 and earlier years. Additionally, the likelihood of tirzepatide initiation was assessed specifically among non-diabetic patients receiving either semaglutide or tirzepatide in the 2023 cohort. Adjusted odds ratios (aOR) and 95% confidence intervals (CIs) were reported. All statistical tests were two-tailed, with a significance level of p < 0.05. Analyses were performed using Python 3 (Python Software Foundation, US).

## Results

A cohort of 319,949 adults eligible for AOMs between January 1, 2015, and January 31, 2024, was identified from OneFlorida+. Of these, 2.0% initiated any AOM, with 0.2% initiating traditional AOMs and 1.8% initiating newer AOMs. Semaglutide accounted for the highest proportion of newer AOM initiations (77.9%), followed by tirzepatide (19.7%) and liraglutide (17.8%). Most individuals initiating newer AOMs were obese (74.9%), female (70.6%), non-Hispanic White (19.1%), aged 40–65 years (60.1%), and privately insured (61.2%). Patient characteristics at baseline are presented in Table 1.

**Table 1.** Characteristics of 2015-2023 OneFlorida+ data trust patients at baseline (n=319949)

|  | **Overall**<br>**eligible cohort**<br>**(n= 319949)** | **Newer AOM**<br>**initiators**<br>**(n= 5835)** | **Traditional**<br>**AOM initiators**<br>**(n= 889)** | **Non-AOM**<br>**initiators**<br>**(n= 313487)** |
| --- | --- | --- | --- | --- |
|  | n (%) | n (%) | n (%) | n (%) |
| **Demographics** |  |  |  |  |
| **Age** |  |  |  |  |
| 18-40 | 98640 (30.8) | 1672 (28.7) | 391 (44.0) | 96694 (30.8) |
| 40-65 | 144218 (45.1) | 3507 (60.1) | 462 (52.0) | 140386 (44.8) |
| ≥65 | 77091 (24.1) | 656 (11.2) | 36 (4.1) | 76407 (24.37) |
| **Sex** |  |  |  |  |
| Female | 193617 (60.5) | 4117 (70.6) | 767 (86.3) | 188959 (60.3) |
| Male | 126318 (39.5) | 1718 (29.4) | 122 (13.7) | 124514 (39.7) |
| **Race and ethnicity** |  |  |  |  |
| White | 110727 (34.6) | 2199 (37.7) | 309 (34.8) | 108306 (34.6) |
| Black | 54508 (17.0) | 966 (16.6) | 149 (16.8) | 53436 (17.1) |
| Hispanic | 66011 (20.6) | 1113 (19.1) | 200 (22.5) | 64765 (20.7) |
| Other | 88703 (27.7) | 1557 (26.7) | 231 (26.0) | 86980 (27.8) |
| **Insurance coverage** |  |  |  |  |
| Private | 172818 (54.0) | 3573 (61.2) | 592 (66.6) | 168837 (53.9) |
| Medicaid | 35207 (11.0) | 519 (8.9) | 84 (9.5) | 34623 (11.0) |
| Medicare | 54350 (17.0) | 674 (11.6) | 40 (4.5) | 53646 (17.1) |
| Uninsured | 60629 (19.0) | 772 (13.2) | 121 (13.6) | 59774 (19.1) |
| Other* | 273798 (85.6) | 5068 (86.9) | 815 (91.7) | 268157 (85.5) |
| **Area deprivation index** |  |  |  |  |
| Decile: 1-2 | 36011 (11.3) | 173 (3.0) | 45 (5.1) | 18454 (5.9) |
| Decile: 3-8 | 137600 (43.0) | 1781 (30.5) | 275 (30.9) | 89609 (28.6) |
| Decile: 9-10 | 60992 (19.1) | 1267 (21.7) | 153 (17.2) | 47768 (15.2) |
| Missing | 85346 (26.7) | 1556 (26.7) | 165 (26.3) | 83625 (26.7) |
| **Index year** |  |  |  |  |
| 2015-2019 | 214510 (67.1) | 4089 (70.1) | 748 (84.1) | 209889 (67.0) |
| 2020-2023 | 105439 (33.0) | 1746 (29.9) | 141 (15.9) | 103598 (33.1) |
| **Comorbidities and risk factors** |  |  |  |  |
| **BMI category** |  |  |  |  |
| Overweight | 103445 (32.3) | 1464 (25.1) | 186 (20.9) | 101852 (32.5) |
| Obese | 216504 (67.7) | 4371 (74.9) | 703 (79.1) | 211635 (67.5) |
| **Coronary artery disease** |  |  |  |  |
| Yes | 12884 (4.0) | 133 (2.3) | 6 (0.7) | 12748 (4.1) |
| **Heart failure** |  |  |  |  |
| Yes | 5546 (1.7) | 39 (0.7) | 1 (0.1) | 5507 (1.8) |
| **Type 2 diabetes** |  |  |  |  |
| Yes | 28083 (8.8) | 942 (16.1) | 24 (2.7) | 27127 (8.7) |
| **Arthritis** |  |  |  |  |
| Yes | 3144 (1.0) | 24 (0.4) | 4 (0.5) | 3117 (1.0) |
| **Chronic obstructive pulmonary disease** |  |  |  |  |
| Yes | 11779 (3.7) | 123 (2.1) | 20 (2.5) | 11643 (3.7) |
| **Anxiety** |  |  |  |  |
| Yes | 24818 (7.8) | 465 (8.0) | 86 (9.7) | 24288 (7.8) |
| **Depression** |  |  |  |  |
| Yes | 18507 (5.8) | 306 (5.2) | 52 (5.9) | 18159 (5.8) |
| **Smoking status** |  |  |  |  |
| Never smoker | 38531 (12.0) | 632 (10.8) | 168 (18.9) | 37780 (12.1) |
| Past smoker | 9608 (3.0) | 125 (2.1) | 18 (2.0) | 9470 (3.0) |
| Current smoker | 4093 (1.3) | 19 (0.3) | 1 (0.1) | 4074 (1.3) |
| **Charlson comorbidity index score** |  |  |  |  |
| 0 | 293785 (91.8) | 5390 (92.4) | 860 (96.7) | 287787 (91.8) |
| 1 | 18124 (5.7) | 297 (5.1) | 21 (2.4) | 17813 (5.7) |
| 2+ | 8040 (2.5) | 148 (2.5) | 8 (0.9) | 7887 (2.5) |
| **Medication use** |  |  |  |  |
| **Antihypertensives** |  |  |  |  |
| Yes | 46801 (14.6) | 1232 (21.1) | 95 (10.7) | 45502 (14.5) |
| **Statins** |  |  |  |  |
| Yes | 27847 (8.7) | 771 (13.2) | 44 (5.0) | 27045 (8.6) |
| **Metformin** |  |  |  |  |
| Yes | 8502 (2.7) | 715 (12.3) | 18 (2.0) | 7776 (2.5) |
| **Other glucose-lowering drugs** |  |  |  |  |
| Yes | 4728 (1.5) | 446 (7.6) | 2 (0.2) | 4282 (1.4) |
| **Antidepressants (Excludes bupropion)** |  |  |  |  |
| Yes | 21995 (6.9) | 410 (7.0) | 69 (7.8) | 21531 (6.9) |
| **Opioids** |  |  |  |  |
| Yes | 89813 (28.1) | 1138 (19.5) | 183 (20.6) | 88538 (28.2) |
| **Provider specialty** |  |  |  |  |
| Cardiology | 15668 (4.9) | 248 (4.3) | 30 (3.4) | 15401 (4.9) |
| Endocrinology | 4928 (1.5) | 261 (4.5) | 23 (2.6) | 4655 (1.5) |
| Nurse practitioner and physician assistant | 65935 (20.6) | 1940 (33.3) | 356 (40.0) | 63757 (20.3) |
| Primary care | 100770 (31.5) | 2495 (42.8) | 453 (51.0) | 97948 (31.2) |
| **Healthcare utilization** |  |  |  |  |
| **Emergency Visits** |  |  |  |  |
| 0 | 174909 (54.7) | 4399 (75.39) | 723 (81.3) | 170002 (54.2) |
| 1+ | 145040 (45.3) | 1436 (24.61) | 166 (18.7) | 143485 (45.8) |
| **Hospitalizations** |  |  |  |  |
| 0 | 269526 (84.2) | 5432 (93.09) | 850 (95.6) | 263497 (84.1) |
| 1+ | 50423 (15.8) | 403 (6.91) | 39 (4.4) | 49990 (16.0) |
| **Outpatient visits** |  |  |  |  |
| 0 | 71827 (22.5) | 751 (12.87) | 74 (8.3) | 71024 (22.7) |
| 1-2 | 73745 (23.1) | 1441 (24.7) | 221 (24.9) | 72146 (23.0) |
| 3+ | 174377 (54.5) | 3643 (62.43) | 594 (66.8) | 170317 (54.3) |

Figure 1 illustrates the proportion of newer AOMs initiated by calendar year from 2015 to 2023. Liraglutide initiation remained relatively stable, starting at 1.5% in 2015, peaking at 5.1% in 2018, and gradually declining to 3.8% by 2023. Semaglutide showed a sharp increase beginning in 2018, rising from 1.4% to 69.1% by 2023. Tirzepatide, introduced in 2022, rapidly grew from 6.5% in its first year to 25.2% in 2023.

**Figure 1.**
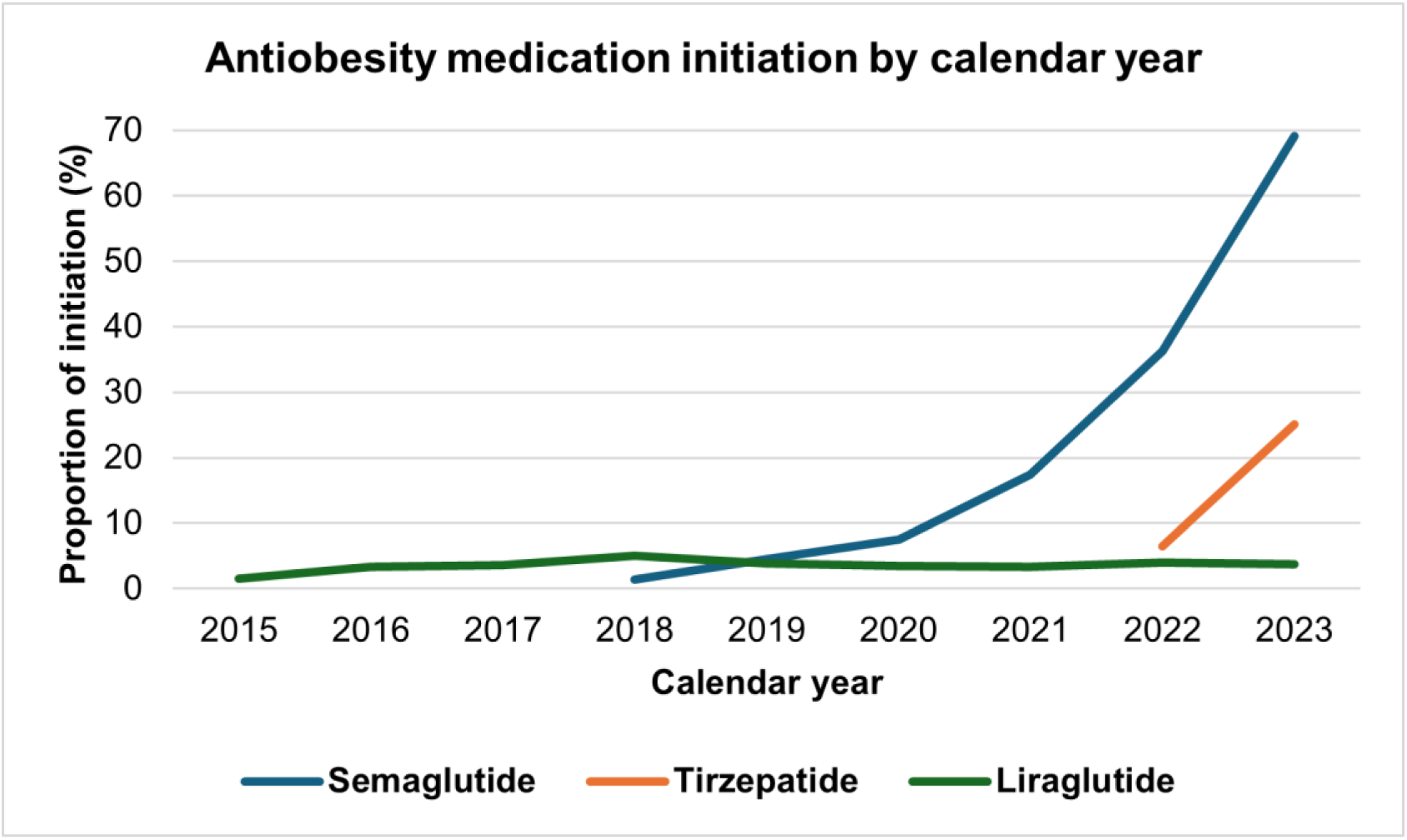
Trends of newer AOM initiation by calendar year

Figures 2a–2d illustrate the proportion of newer AOM initiation in 2022 and 2023 by ZIP code areas in Florida. Darker colors indicate higher uptake; gray represents missing data. Overall, higher AOM uptake was seen in Central, North, and Southeast Florida, with lower uptake in the Southwest, Northwest, and Panhandle. Semaglutide had higher uptake in Central and North Florida and parts of Southeast Florida. Liraglutide showed higher uptake in scattered Central and Southeast counties, with lower uptake in the Panhandle and North Florida. Tirzepatide uptake was higher in select Central and Southeast counties and lower in the Panhandle and Southwest. Supplementary Figure S1 presents ZIP code-level AOM uptake for all study years.

**Figure 2.**
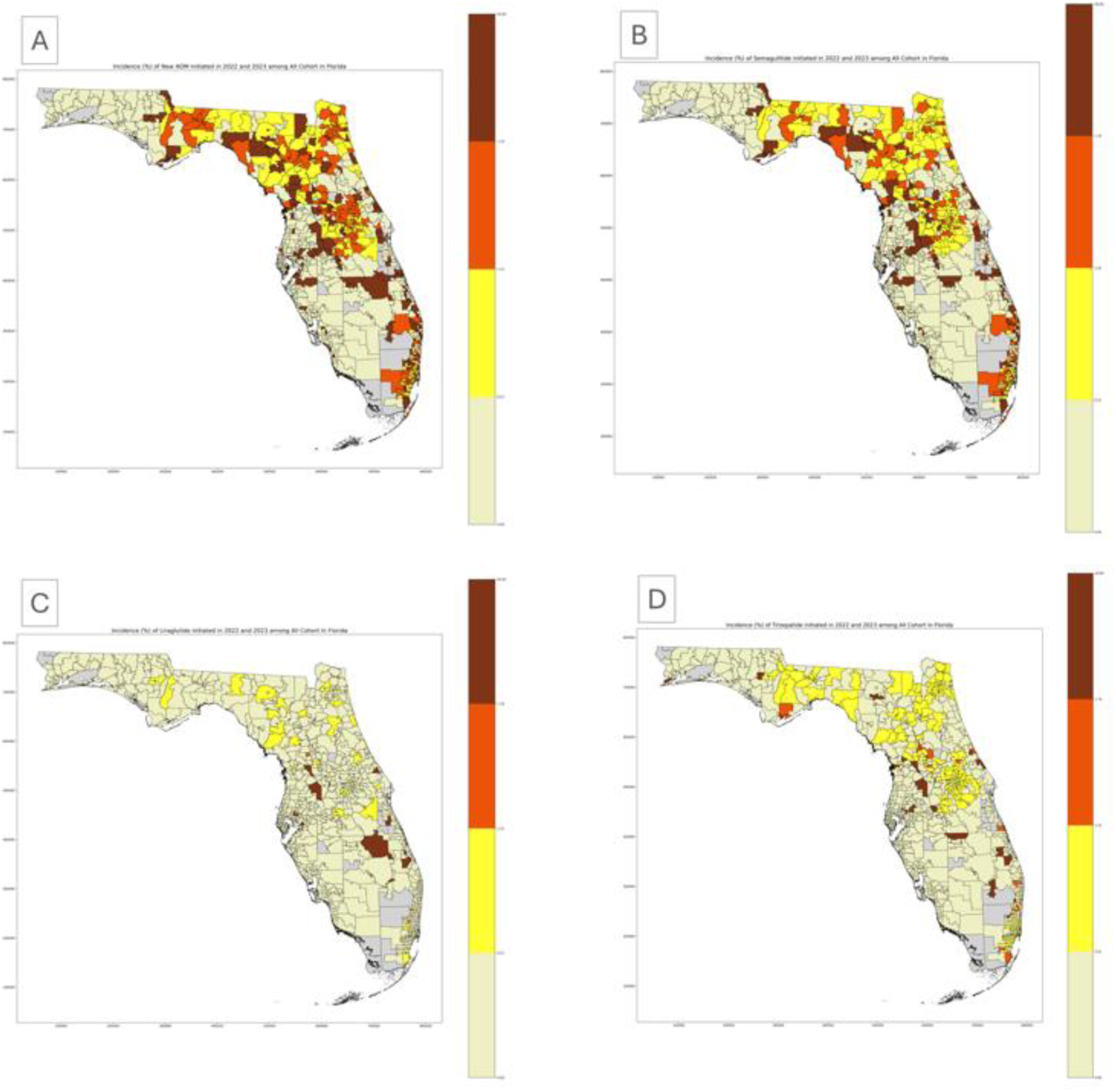
Incidence proportion of initiating overall newer AOMs and each individual drug among the eligible cohort in year 2022 and 2023, stratified by five-digit ZIP codes in Florida. (A) Overall newer AOM initiation, (B) semaglutide initiation, (C) liraglutide initiation, and (D) tirzepatide initiation.

### Factors associated with newer AOM initiation among the eligible cohort

Sociodemographic factors—including age, gender, race or ethnicity, and insurance status—were significantly associated with newer AOM initiation (Table 2). Higher odds were found among females compared to males (aOR 1.60 [1.51, 1.70]) and individuals aged 40–65 years compared to 18–40 years (1.21 [1.13, 1.29]). Lower odds were found in individuals aged ≥65 years compared to 18–40 years (0.38 [0.34, 0.43]), non-Hispanic Black (0.87 [0.80, 0.94]), Hispanic (0.84 [0.78, 0.91]), and other race or ethnic groups (0.89 [0.83, 0.95]) compared to the non-Hispanic White group. Those with Medicaid (0.69 [0.63, 0.76]), Medicare (0.86 [0.78, 0.95]), other insurance (0.90 [0.83, 0.98]), or no insurance (0.81 [0.74, 0.87]) had lower odds compared to private insurance.

**Table 2.** Factors associated with initiating newer AOMs (n= 5835) among all eligible cohort (n= 319949)

| Variable | aOR | 95% CI | p-value |
| --- | --- | --- | --- |
| **Demographics** |  |  |  |
| **Age (ref= 18-40)** |  |  |  |
| 40-65 | 1.21 | 1.13, 1.29 | <0.001 |
| ≥65 | 0.38 | 0.34, 0.43 | <0.001 |
| **Sex (ref= male)** |  |  |  |
| Female | 1.60 | 1.51, 1.70 | <0.001 |
| **Race and ethnicity (ref= White)** |  |  |  |
| Black | 0.87 | 0.8, 0.94 | <0.001 |
| Hispanic | 0.84 | 0.78, 0.91 | <0.001 |
| Other | 0.89 | 0.83, 0.95 | 0.001 |
| **Insurance coverage (ref= Private)** |  |  |  |
| Medicaid | 0.69 | 0.63, 0.76 | <0.001 |
| Medicare | 0.86 | 0.78, 0.95 | 0.002 |
| Uninsured | 0.81 | 0.74, 0.87 | <0.001 |
| Other | 0.90 | 0.83, 0.98 | 0.014 |
| **Area Deprivation index (ref= 1-2)** |  |  |  |
| Decile: 3-8 | 1.03 | 0.94, 1.12 | 0.548 |
| Decile: 9-10 | 0.98 | 0.89, 1.09 | 0.753 |
| Missing | 1.01 | 0.92, 1.11 | 0.899 |
| **Comorbidities and risk factors** |  |  |  |
| **BMI category (ref= overweight)** |  |  |  |
| Obesity | 1.66 | 1.55, 1.77 | <0.001 |
| **Coronary artery disease (ref= no)** |  |  |  |
| Yes | 0.82 | 0.68, 0.99 | 0.039 |
| **Heart failure (ref= no)** |  |  |  |
| Yes | 0.55 | 0.39, 0.77 | <0.001 |
| **Type 2 diabetes (ref= no)** |  |  |  |
| Yes | 2.19 | 1.99, 2.40 | <0.001 |
| **Arthritis (ref= no)** |  |  |  |
| Yes | 0.49 | 0.33, 0.74 | 0.001 |
| **Chronic obstructive pulmonary disease (ref= no)** |  |  |  |
| Yes | 0.84 | 0.69, 1.01 | 0.066 |
| **Anxiety (ref= no)** |  |  |  |
| Yes | 1.13 | 1.01, 1.26 | 0.037 |
| **Depression (ref= no)** |  |  |  |
| Yes | 0.96 | 0.84, 1.10 | 0.544 |
| **Smoking (ref= never smoker)** |  |  |  |
| Current smoker | 0.37 | 0.24, 0.58 | <0.001 |
| Former smoker | 0.81 | 0.67, 0.97 | 0.023 |
| **Charlson comorbidity index score (ref= 0)** |  |  |  |
| 1 | 0.84 | 0.73, 0.95 | **0.007** |
| 2+ | 1.01 | 0.84, 1.21 | 0.922 |
| **Medication use** |  |  |  |
| **Antihypertensives (ref= no)** |  |  |  |
| Yes | 1.39 | 1.25, 1.55 | **<0.001** |
| **Statins (ref= no)** |  |  |  |
| Yes | 0.95 | 0.84, 1.07 | 0.39 |
| **Metformin (ref= no)** |  |  |  |
| Yes | 2.94 | 2.60, 3.31 | **<0.001** |
| **Other glucose-lowering drugs (ref= no)** |  |  |  |
| Yes | 2.22 | 1.93, 2.56 | **<0.001** |
| **Antidepressants (Excludes bupropion) (ref= no)** |  |  |  |
| Yes | 1.11 | 0.99, 1.24 | 0.081 |
| **Opioids (ref= no)** |  |  |  |
| Yes | 0.83 | 0.77, 0.89 | **<0.001** |
| **Provider specialty (ref= primary care)** |  |  |  |
| Cardiology | 1.00 | 0.87, 1.14 | 0.955 |
| Endocrinology | 1.92 | 1.67, 2.20 | **<0.001** |
| Nurse practitioner and physician assistant | 1.60 | 1.50, 1.71 | **<0.001** |
| **Healthcare utilization** |  |  |  |
| **ER visits (ref= 0)** |  |  |  |
| 1+ | 0.48 | 0.45, 0.51 | **<0.001** |
| **Hospitalization (ref= 0)** |  |  |  |
| 1+ | 0.54 | 0.48, 0.61 | **<0.001** |
| **Outpatient visits (ref= 0)** |  |  |  |
| 1-2 | 1.31 | 1.20, 1.44 | **<0.001** |
| 3+ | 1.26 | 1.15, 1.38 | **<0.001** |

Abbreviations: aOR, adjusted odds ratio, CI, confidence interval Bold values indicate significance at alpha= 0.05 \*Other insurance coverage indicates those with more than one type of insurance coverage

Among clinical variables, higher odds of initiating newer AOMs were observed among individuals with obesity (1.66, [1.55, 1.77]) compared to those who were overweight, as well as those with T2D (2.19, [1.99, 2.40]) or anxiety (1.13, [1.01, 1.26]). Similarly, higher odds were associated with the use of metformin (2.94, [2.60, 3.31]), other glucose-lowering agents (2.22, [1.93, 2.56]), and antihypertensives (1.39, [1.25, 1.55]). In contrast, lower odds were observed among individuals with coronary artery disease (0.82, [0.68, 0.99]), heart failure (0.55, [0.39, 0.77]), or arthritis (0.49, [0.33, 0.74]). Current smokers (0.37, [0.24, 0.58]) and former smokers (0.81, [0.67, 0.97]) had lower odds compared to never smokers. Lower odds were also seen among opioid users (0.83, [0.77, 0.89]) and those with a Charlson Comorbidity Index score of 1 (0.84, [0.73, 0.95]) compared to 0.

Regarding healthcare utilization, individuals with 1-2 (1.31, [1.20, 1.44]) or ≥3 outpatient visits (1.26, [1.15, 1.38]) had higher odds of AOM initiation compared to none. Consulting an endocrinologist (1.92, [1.67, 2.20]) or a specialized nurse practitioner/physician assistant (1.60, [1.50, 1.71]) was also associated with higher odds compared to primary care providers. Lower odds were associated to ≥1 emergency room visits (0.48, [0.45, 0.51]) or hospitalizations (0.54, [0.48, 0.61]) compared to none.

### Factors associated with newer AOM initiation stratified by diabetes status

In individuals with and without T2D, higher odds of initiating newer AOMs were observed among those with obesity, anxiety, antihypertensive or metformin use, visits to an endocrinologist or specialized nurse/physician assistant, and ≥1 outpatient visits. Lower odds were noted among individuals aged 65 or older, current smokers, and those with hospitalizations or emergency room visits (Tables S2 and S3).

Among non-diabetic individuals, higher odds were seen in females, those aged 40–65 years, and antidepressant users. Lower odds were associated with former smoking, heart failure, coronary artery disease, arthritis, opioid use, minority racial or ethnic groups, and public or no insurance (Table S2). For diabetic individuals, lower odds were observed among those aged 65 or older, Medicaid insurance, heart failure, and current smoking, (Table S3).

### Factors associated with newer AOM initiation stratified by year of initiation

Findings stratified by initiation in 2023 versus prior years were similar. Odds were higher among those with obesity, females, middle age, users of metformin, antihypertensives, or antidepressants, those who visited an endocrinologist or specialized nurse/physician assistant, and ≥1 outpatient visits. Lower odds were observed among minority populations, those with public or no insurance, individuals aged 65 or older, opioid users, current smokers, those with a Charlson Comorbidity Index score of 1, and those with ≥1 hospitalizations or emergency room visits (Tables S4 and S5). In 2023, additional factors associated with lower odds included anxiety, the use of other glucose-lowering medications, coronary artery disease, and former smoking (Table S5).

### Factors associated with tirzepatide initiation

Among non-diabetic patients receiving either semaglutide or tirzepatide in the 2023 cohort, patients using statins were less likely to initiate tirzepatide (0.54, [0.33, 0.89]). Similarly, non-Hispanic Black (0.65, [0.49, 0.87]) and Hispanic patients (0.75, [0.57, 0.98]) had lower odds of initiation of tirzepatide compared to non-Hispanic White patients. Healthcare utilization also played a role, with patients who had one or more emergency room visits (0.76, [0.60, 0.96]) or 1–2 outpatient visits (0.68, [0.51, 0.92]) being less likely to initiate tirzepatide compared to those with no visits (Table S6)

## Discussion

In a cohort of real-world populations eligible for AOM treatment, we observed that only 2.0% and 1.8% initiated any or a newer AOM between January 1, 2015, and January 31, 2024. We identified significant disparities in initiating newer AOMs. For example, racial and ethnic minority groups were less likely to initiate a newer AOM; uninsured people or public insurance enrollees were less likely to use a newer AOM compared to private insurance enrollees. We also found that health care use, e.g., patients having regular outpatient visits and seeing an endocrinologist, were more likely to be prescribed newer AOMs. Other factors influencing initiation included age, sex, smoking status, comorbidities and medication use. Greater odds of initiation were associated with being female, middle-aged, having T2D, and using glucose-lowering drugs, or antihypertensive medications.

Our results align with previous research indicating disparities in the use of AOMs across racial and ethnic groups in the US (11, 17). Gasoyan et al. found significant disparities in access to AOMs, both in terms of prescribing and filling prescriptions. Specifically, Black, Hispanic, and other racial or ethnic groups (compared to Whites) had lower odds of receiving a newer prescription for AOMs, and Hispanics had lower odds of having the prescription filled (11).

Similarly, a cross-sectional analysis of the 2015-2020 National Health and Nutrition Examination Survey reported that many Americans eligible for semaglutide for obesity were likely unable to afford it, with Black and Hispanic adults having less access than White individuals; this difference was attributable to differences in insurance status, having a usual source of care, family income, and education (17). Our subgroup analyses, which included patients without diabetes and cohorts from both 2023 and earlier years, further corroborated these findings, showing that belonging to a minority racial or ethnic group was consistently associated with lower odds of receiving AOMs. Both racial and weight-related biases are documented to influence healthcare interactions (18–20). These biases can impact how patients communicate with healthcare providers and their willingness to seek treatment for obesity, potentially leading to underutilization of effective treatments like newer AOM (18–20). More research is needed to understand intrinsic (e.g., perceptions of clinicians or patients), and extrinsic factors (e.g., structural or external barriers) that contribute to racial and ethnic disparities in the treatment of obesity.

Insurance coverage is a barrier to initiating AOMs, as evidenced by our findings and those of Gasyon et al. (11). Individuals covered by Medicaid, Medicare, other types of insurance, or no insurance have lower odds of initiating AOMs compared to those with private insurance.

Unfortunately, for an obesity indication, most state Medicaid programs and Medicare Part D plans do not cover AOMs, including Florida (21; 22). Among those with private insurance, coverage for AOMs varies by carrier and often involves strict preauthorization (23). In contrast, access to GLP-1 RAs to treat T2D has better coverage. Our subgroup analyses showed that for individuals with T2D, only Medicaid enrollees had significantly lower odds of initiating AOMs. For those without diabetes, significant disparities exist, with all public insurance enrollees (Medicaid and Medicare) having lower odds of initiating AOMs. When Medicare Part D was established, traditional AOMs were excluded from coverage due to unfavorable safety profiles (6). However, newer AOMs have demonstrated significant weight reduction with improved safety profiles (7; 8). In response, promising changes are underway. More recently, in 2024, the Biden-Harris Administration proposed expanding Medicare and Medicaid coverage for AOMs to improve accessibility and affordability. This expansion is estimated to reduce Medicare enrollees’ out-of-pocket costs by up to 95% and benefit 4 million Medicaid enrollees (24).

Provider specialty was significantly associated with initiating newer AOMs. Our findings showed that patients who visited an endocrinologist or a specialized nurse practitioner/physician assistant were more likely to initiate AOMs compared to those who visited a primary care provider. Similarly, Thomas et al. found that endocrinologists had the highest prevalence of prescribing newer AOMs, reflecting their expertise in this area (27). However, projections from 2003 and 2014 reveal that the endocrinology workforce is insufficient to meet current and future demand to significantly impact the obesity epidemic (28). There is also substantial US nationwide variations in geographic accessibility to endocrinologists at both county and state levels, as well as disparities in access to an endocrinologist within a reasonable driving distance, by urban/rural status and by age (29). The unequal geographic distribution, including shortages, of the endocrine workforce in the US, highlight areas with limited access to these specialists (29), which may continue to negatively impact AOM use for those who need them. A potential solution is the use of telehealth to improve access to endocrinologists at a lower cost and with reduced travel time (30). Additionally, promoting the growth of the endocrinology profession by creating systematic and institutional incentives for endocrinology trainees, such as funding opportunities or loan repayment, could help address the workforce shortage (31).

The association between clinical characteristics and newer AOM initiation varies across studies. In our study, individuals with T2D and those using metformin, glucose-lowering, or anti-hypertensive medications were more likely to initiate newer AOMs. Conversely, individuals with coronary artery disease, heart failure, arthritis, or higher Charlson comorbidity scores were less likely. Another study found that higher Charlson comorbidity scores, myocardial infarction, or heart failure lowered the odds of both receiving and filling an AOM prescription. However, patients with T2D had lower odds of receiving an AOM prescription but higher odds of filling one (11). The authors proposed that patients with diabetes might be prescribed liraglutide or semaglutide primarily for glucose control with added weight loss benefits (11). A retrospective study investigating obesity treatment modalities reported that weight reduction procedures or surgeries were more common among individuals with severe obesity and higher comorbidity burdens, while AOM use was associated with cardiovascular risk factors (32). Unlike our study, which focuses exclusively on newer AOMs, these studies included traditional AOMs in their analyses. The difference in safety profiles and the larger body of evidence supporting surgical outcomes compared to traditional AOMs may have influenced clinical preferences (33–35). Our findings emphasize the need for increased awareness of newer AOMs’ cardiometabolic benefits, which could shift prescribing patterns over time. Further research is essential to update treatment guidelines for obesity, particularly for newer AOMs.

Our findings carry significant public health and clinical implications, offering real-world data on newer AOM uptake, including trends over time and geographic variation across Florida. We identified lower initiation rates among racial and ethnic minorities, the uninsured, and those with limited healthcare access, revealing systemic barriers to obesity treatment. These findings support designing targeted interventions and policies to improve AOM equity and ultimately clinical outcomes. We propose strategies to increase AOM uptake. These include advocating for policy reforms to expand insurance coverage and ease prior authorization requirements. To improve access to obesity specialists, we recommend fostering the growth of the endocrinology field through targeted incentives for trainees and expanding telehealth services. Finally, previous research revealed clinician and patient concerns about AOM side effects and limited awareness of their benefits (36–38). Prioritizing outreach efforts is crucial to educate both groups about newer AOMs, including improving understanding of obesity science, updating obesity management guidelines, and recognizing obesity as a chronic disease.

Several limitations of this study should be noted. First, data on patients’ income or educational level were not available and, therefore, could not be accounted for. However, the ADI was used as a close proxy, but because it is multifaceted, it may not fully capture the individual-level socioeconomic differences that could impact health behaviors and access to care, potentially leading to residual confounding in our analysis. Second, the use of secondary data from the OneFlorida+ database limited our ability to capture certain latent variables, such as patient or clinician perceptions and beliefs, that could have influenced the initiation of newer AOMs. Third, while a strength of our study is its focus on newer AOMs, including tirzepatide, it was difficult to directly compare our findings with previous studies, which primarily included traditional AOMs. When newer AOMs were included, they typically involved semaglutide, liraglutide, or both, but none included tirzepatide. Finally, because our focus was on investigating the initiation of newer AOMs, patients with any AOM use at baseline were excluded. As a result, we were unable to assess whether prior use of traditional AOMs influenced the decision to initiate a newer AOM.

In conclusion, the initiation of newer AOMs remains low among overweight and obese adults in the US but has notably increased since 2022. Our findings underscore significant disparities in obesity care, with racial and ethnic minority groups and individuals without private insurance being less likely to access these treatments. Addressing inequities in AOM access is essential to mitigate disparities in obesity outcomes.

## Funding and assistance

NIH/NIDDK R01DK133465

## Author contributions and guarantor statement

R. M. R, Y.A.L, and J.G. were involved in the conception, design, and conduct of the study and the analysis and interpretation of the results. R.M.R. wrote the first draft of the manuscript, and all authors reviewed and approved the final version of the manuscript. J. G. is the guarantor of this work and, as such, had full access to all the data in the study and takes responsibility for the integrity of the data and the accuracy of the data analysis.

## Data Availability

All data produced in the present study are available upon reasonable request to the authors.

