## Supplementary material for "Trends and Disparities in Newer GLP1 Receptor Agonist Initiation among Real-World Adult Patients Eligible for Obesity Treatment"

**Contents**

**Figure S1**. Incidence proportion of overall newer AOM initiation and individual drugs among the eligible cohort across all years, stratified by five-digit ZIP codes in Florida.

**Table S2.** Factors associated with initiating newer AOMs (n= 4893) among a cohort of patients without type 2 diabetes (n= 291866)

**Table S3.** Factors associated with initiating newer AOMs (n= 942) among a cohort of patients with type 2 diabetes (n= 28083)

**Table S4.** Factors associated with initiating newer AOMs prior to year 2023 (n= 2963) among a cohort of new AOM users (n= 5835) and non AOM users (n= 313487).

**Table S5.** Factors associated with initiating newer AOMs in year 2023 and after (n= 2872) among a cohort of new AOM users (n= 5835) and non AOM users (n= 313487).

**Table S6.** Factors associated with initiating tirzepatide (n= 712) among a cohort of patients without type 2 diabetes, receiving semaglutide (n= 1765) or tirzepatide (n= 712) in 2023.

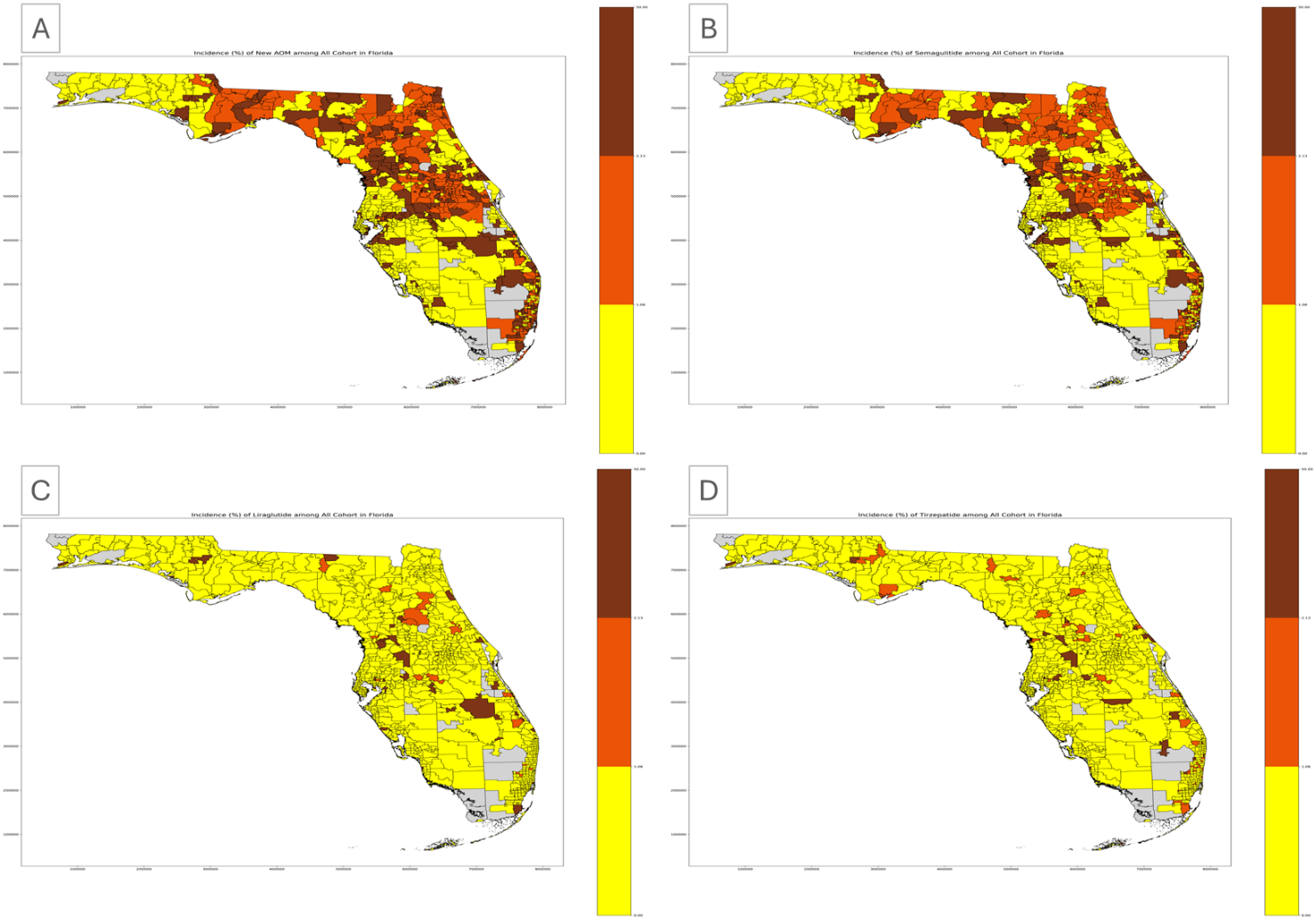

Figure S1. Incidence proportion of overall newer AOM initiation and individual drugs among the eligible cohort across all years, stratified by five-digit ZIP codes in Florida. (A) Overall newer AOM initiation, (B) semaglutide initiation, (C) liraglutide initiation, and (D) tirzepatide initiation.

**Table S1.** Factors associated with initiating any AOMs (n= 6462) among all eligible cohort (n= 319949)

| **Variable** | **aOR** | **95% CI** | **p-value** |
| --- | --- | --- | --- |
| **Demographics** |  |  |  |
| **Age (ref= 18-40)** |  |  |  |
| 40-65 | 1.15 | 1.08, 1.22 | **<0.001** |
| ≥65 | 0.36 | 0.32, 0.40 | **<0.001** |
| **Sex (ref= male)** |  |  |  |
| Female | 1.71 | 1.61, 1.81 | **<0.001** |
| **Race and ethnicity (ref= White)** |  |  |  |
| Black | 0.88 | 0.81, 0.94 | **0.001** |
| Hispanic | 0.86 | 0.80, 0.92 | **<0.001** |
| Other | 0.90 | 0.84, 0.96 | **0.001** |
| **Insurance coverage (ref= Private)** |  |  |  |
| Medicaid | 0.68 | 0.62, 0.75 | **<0.001** |
| Medicare | 0.83 | 0.76, 0.92 | **<0.001** |
| Uninsured | 0.81 | 0.75, 0.87 | **<0.001** |
| Other | 0.94 | 0.87, 1.01 | **0.096** |
| **Area Deprivation index (ref= 1-2)** |  |  |  |
| Decile: 3-8 | 1.03 | 0.95, 1.12 | 0.468 |
| Decile: 9-10 | 0.99 | 0.90, 1.09 | 0.906 |
| Missing | 1.01 | 0.92, 1.10 | 0.828 |
| **Comorbidities and risk factors** |  |  |  |
| **BMI category (ref= overweight)** |  |  |  |
| Obesity | 1.67 | 1.57, 1.78 | **<0.001** |
| **Coronary artery disease (ref= no)** |  |  |  |
| Yes | 0.81 | 0.67, 0.98 | **0.026** |
| **Heart failure (ref= no)** |  |  |  |
| Yes | 0.53 | 0.38, 0.74 | **<0.001** |
| **Type 2 diabetes (ref= no)** |  |  |  |
| Yes | 2.05 | 1.87, 2.25 | **<0.001** |
| **Arthritis (ref= no)** |  |  |  |
| Yes | 0.50 | 0.34, 0.74 | **<0.001** |
| **Chronic obstructive pulmonary disease (ref= no)** |  |  |  |
| Yes | 0.86 | 0.72, 1.03 | 0.099 |
| **Anxiety (ref= no)** |  |  |  |
| Yes | 1.14 | 1.03, 1.27 | 0.016 |
| **Depression (ref= no)** |  |  |  |
| Yes | 0.98 | 0.86, 1.11 | 0.708 |
| **Smoking (ref= never smoker)** |  |  |  |
| Current smoker | 0.34 | 0.21, 0.53 | **<0.001** |
| Former smoker | 0.81 | 0.68, 0.97 | **0.019** |
| **Charlson comorbidity index score (ref= 0)** |  |  |  |
| 1 | 0.85 | 0.74, 0.96 | **0.011** |
| 2+ | 1.02 | 0.85, 1.21 | 0.842 |
| **Medication use** |  |  |  |
| **Antihypertensives (ref= no)** |  |  |  |
| Yes | 1.37 | 1.24, 1.52 | **<0.001** |
| **Statins (ref= no)** |  |  |  |
| Yes | 0.94 | 0.84, 1.06 | 0.341 |
| **Metformin (ref= no)** |  |  |  |
| Yes | 2.84 | 2.53, 3.20 | **<0.001** |
| **Other glucose-lowering drugs (ref= no)** |  |  |  |
| Yes | 2.17 | 1.89, 2.50 | **<0.001** |
| **Antidepressants (Excludes bupropion) (ref= no)** |  |  |  |
| Yes | 1.14 | 1.02, 1.27 | **0.016** |
| **Opioids (ref= no)** |  |  |  |
| Yes | 0.87 | 0.81, 0.93 | **<0.001** |
| **Provider specialty (ref= primary care)** |  |  |  |
| Cardiology | 1.00 | 0.87, 1.13 | 0.951 |
| Endocrinology | 1.85 | 1.61, 2.11 | **<0.001** |
| Nurse practitioner and physician assistant | 1.61 | 1.52, 1.71 | **<0.001** |
| **Healthcare utilization** |  |  |  |
| **ER visits (ref= 0)** |  |  |  |
| 1+ | 0.46 | 0.43, 0.49 | **<0.001** |
| **Hospitalization (ref= 0)** |  |  |  |
| 1+ | 0.53 | 0.48, 0.59 | **<0.001** |
| **Outpatient visits (ref= 0)** |  |  |  |
| 1-2 | 1.35 | 1.24, 1.48 | **<0.001** |
| 3+ | 1.31 | 1.20, 1.43 | **<0.001** |
| Abbreviations: aOR, adjusted odds ratio, CI, confidence interval  Bold values indicate significance at alpha= 0.05  *Other insurance coverage indicates those with more than one type of insurance coverage | | | |

**Table S2.** Factors associated with initiating newer AOMs (n= 4893) among a cohort of patients without type 2 diabetes (n= 291866)

| **Variable** | **aOR** | **95% CI** | | **p-value** |
| --- | --- | --- | --- | --- |
| **Demographics** |  |  |  |  |
| **Age (ref= 18-40)** |  |  |  |  |
| 40-65 | 1.25 | 1.17, 1.33 | | **<0.001** |
| ≥65 | 0.41 | 0.36, 0.46 | | **<0.001** |
| **Sex (ref= male)** |  |  |  |  |
| Female | 1.71 | 1.60, 1.83 | | **<0.001** |
| **Race and ethnicity (ref= White)** |  |  |  |  |
| Black | 0.87 | 0.80, 0.95 | | **0.002** |
| Hispanic | 0.82 | 0.75, 0.89 | | **<0.001** |
| Other | 0.90 | 0.84, 0.97 | | **0.004** |
| **Insurance coverage (ref= Private)** | |  |  |  |
| Medicaid | 0.69 | 0.62, 0.76 | | **<0.001** |
| Medicare | 0.83 | 0.74, 0.93 | | **0.002** |
| Uninsured | 0.79 | 0.72, 0.87 | | **<0.001** |
| Other | 0.89 | 0.81, 0.97 | | **0.006** |
| **Area Deprivation index (ref= 1-2)** | |  |  |  |
| Decile: 3-8 | 1.00 | 0.91, 1.10 | | 0.997 |
| Decile: 9-10 | 0.98 | 0.88, 1.09 | | 0.707 |
| Missing | 1.00 | 0.90, 1.11 | | 0.980 |
| **Comorbidities and risk factors** |  |  |  |  |
| **BMI category (ref= overweight)** | |  |  |  |
| Obesity | 1.66 | 1.54, 1.78 | | **<0.001** |
| **Coronary artery disease (ref= no)** | |  |  |  |
| Yes | 0.72 | 0.54, 0.97 | | **0.031** |
| **Heart failure (ref= no)** |  |  |  |  |
| Yes | 0.49 | 0.27, 0.90 | | **0.022** |
| **Arthritis (ref= no)** |  |  |  |  |
| Yes | 0.42 | 0.25, 0.71 | | **0.001** |
| Yes | 0.90 | 0.71, 1.14 | | 0.387 |
| **Anxiety (ref= no)** |  |  |  |  |
| Yes | 1.09 | 0.96, 1.24 | | 0.175 |
| **Depression (ref= no)** |  |  |  |  |
| Yes | 0.97 | 0.83, 1.14 | | 0.733 |
| **Smoking (ref= never smoker)** | |  |  |  |
| Current smoker | 0.43 | 0.27, 0.69 | | **0.001** |
| Former smoker | 0.74 | 0.59, 0.92 | | **0.006** |
| **Charlson comorbidity index score (ref= 0)** |  |  | |  |
| 1 | 0.80 | 0.64, 1.01 | | 0.055 |
| 2+ | 1.01 | 0.71, 1.42 | | 0.965 |
| **Medication use** |  |  |  |  |
| **Antihypertensives (ref= no)** |  |  |  |  |
| Yes | 1.43 | 1.28, 1.61 | | **<0.001** |
| **Statins (ref= no)** |  |  |  |  |
| Yes | 0.95 | 0.82, 1.10 | | 0.456 |
| **Metformin (ref= no)** |  |  |  |  |
| Yes | 4.55 | 3.98, 5.21 | | **<0.001** |
| **Antidepressants (Excludes bupropion) (ref= no)** |  |  |  |  |
| Yes | 1.17 | 1.04, 1.33 | | **0.011** |
| **Opioids (ref= no)** |  |  |  |  |
| Yes | 0.82 | 0.75, 0.89 | | **<0.001** |
| **Provider specialty (ref= primary care)** |  |  |  |  |
| Cardiology | 0.97 | 0.83, 1.13 | | 0.683 |
| Endocrinology | 2.12 | 1.80, 2.50 | | **<0.001** |
| Nurse practitioner and physician assistant | 1.66 | 1.55, 1.78 | | **<0.001** |
| **Healthcare utilization** |  |  |  |  |
| **ER visits (ref= 0)** |  |  |  |  |
| 1+ | 0.46 | 0.43, 0.50 | | **<0.001** |
| **Hospitalization (ref= 0)** |  |  |  |  |
| 1+ | 0.56 | 0.49, 0.64 | | **<0.001** |
| **Outpatient visits (ref= 0)** |  |  |  |  |
| 1-2 | 1.26 | 1.15, 1.40 | | **<0.001** |
| 3+ | 1.18 | 1.07, 1.30 | | **0.001** |
| Abbreviations: aOR, adjusted odds ratio, CI, confidence interval |  |  |  |  |
| Bold values indicate significance at alpha= 0.05 |  | | |  |
| *Other insurance coverage indicates those with more than one type of insurance coverage | | | | |

**Table S3.** Factors associated with initiating newer AOMs (n= 942) among a cohort of patients with type 2 diabetes (n= 28083)

| **Variable** | **aOR** | **95% CI** | | **p-value** |
| --- | --- | --- | --- | --- |
| **Demographics** |  |  |  |  |
| **Age (ref= 18-40)** |  |  |  |  |
| 40-65 | 0.81 | 0.65, 1.02 | | 0.077 |
| ≥65 | 0.25 | 0.19, 0.33 | | **<0.001** |
| **Sex (ref= male)** |  |  |  |  |
| Female | 1.13 | 0.98, 1.29 | | 0.091 |
| **Race and ethnicity (ref= White)** | |  |  |  |
| Black | 0.86 | 0.70, 1.05 | | 0.142 |
| Hispanic | 0.96 | 0.79, 1.16 | | 0.650 |
| Other | 0.96 | 0.76, 1.22 | | 0.757 |
| **Insurance coverage (ref= Private)** | |  |  |  |
| Medicaid | 0.68 | 0.54, 0.86 | | **0.001** |
| Medicare | 0.94 | 0.78, 1.13 | | 0.483 |
| Uninsured | 0.85 | 0.71, 1.02 | | 0.076 |
| Other | 0.87 | 0.72, 1.04 | | 0.118 |
| **Area Deprivation index (ref= 1-2)** | |  |  |  |
| Decile: 3-8 | 1.18 | 0.94, 1.48 | | 0.150 |
| Decile: 9-10 | 1.01 | 0.78, 1.31 | | 0.920 |
| Missing | 1.05 | 0.83, 1.35 | | 0.668 |
| **Comorbidities and risk factors** | |  |  |  |
| **BMI category (ref= overweight)** | |  |  |  |
| Obesity | 1.59 | 1.37, 1.85 | | **<0.001** |
| **Coronary artery disease (ref= no)** | |  |  |  |
| Yes | 0.90 | 0.69, 1.16 | | 0.395 |
| **Heart failure (ref= no)** |  |  |  |  |
| Yes | 0.59 | 0.39, 0.90 | | **0.013** |
| **Arthritis (ref= no)** |  |  |  |  |
| Yes | 0.65 | 0.34, 1.24 | | 0.191 |
| **Chronic obstructive pulmonary disease (ref= no)** | | |  |  |
| Yes | 0.77 | 0.56, 1.05 | | 0.095 |
| **Anxiety (ref= no)** |  |  |  |  |
| Yes | 1.30 | 1.01, 1.65 | | **0.038** |
| **Depression (ref= no)** |  |  |  |  |
| Yes | 0.92 | 0.70, 1.21 | | 0.556 |
| **Smoking (ref= never smoker)** | |  |  |  |
| Current smoker | 0.10 | 0.01, 0.69 | | **0.020** |
| Former smoker | 1.03 | 0.74, 1.43 | | 0.867 |
| **Charlson comorbidity index score (ref= 0)** | |  |  |  |
| 1 | 0.88 | 0.74, 1.04 | | 0.143 |
| 2+ | 1.00 | 0.81, 1.24 | | 0.967 |
| **Medication use** |  |  |  |  |
| **Antihypertensives (ref= no)** | |  |  |  |
| Yes | 1.38 | 1.08, 1.76 | | **0.009** |
| **Statins (ref= no)** |  |  |  |  |
| Yes | 1.18 | 0.93, 1.50 | | 0.171 |
| **Metformin (ref= no)** |  |  |  |  |
| Yes | 3.09 | 2.60, 3.68 | | **<0.001** |
| **Antidepressants (Excludes bupropion) (ref= no)** | | |  |  |
| Yes | 0.91 | 0.70, 1.19 | | 0.509 |
| **Opioids (ref= no)** |  |  |  |  |
| Yes | 0.90 | 0.76, 1.06 | | 0.213 |
| **Provider specialty (ref= primary care)** | |  |  |  |
| Cardiology | 1.07 | 0.82, 1.41 | | 0.609 |
| Endocrinology | 1.64 | 1.28, 2.11 | | **<0.001** |
| Nurse practitioner and physician assistant | 1.31 | 1.11, 1.55 | | **0.002** |
| **Healthcare utilization** |  |  |  |  |
| **ER visits (ref= 0)** |  |  |  |  |
| 1+ | 0.58 | 0.49, 0.68 | | **<0.001** |
| **Hospitalization (ref= 0)** |  |  |  |  |
| 1+ | 0.48 | 0.38, 0.60 | | **<0.001** |
| **Outpatient visits (ref= 0)** |  |  |  |  |
| 1-2 | 1.74 | 1.30, 2.33 | | **<0.001** |
| 3+ | 1.98 | 1.51, 2.59 | | **<0.001** |
| Abbreviations: aOR, adjusted odds ratio, CI, confidence interval | | | |  |
| Bold values indicate significance at alpha= 0.05 | | |  |  |
| *Other insurance coverage indicates those with more than one type of insurance coverage | | | | |

**Table S4.** Factors associated with initiating newer AOMs prior to year 2023 (n= 2963) among a cohort of new AOM users (n= 5835) and eligible non AOM users (n= 313487).

| **Variable** | **aOR** | **95% CI** | | **p-value** |
| --- | --- | --- | --- | --- |
| **Demographics** |  |  |  |  |
| **Age (ref= 18-40)** |  |  |  |  |
| 40-65 | 1.35 | 1.23, 1.48 | | **<0.001** |
| ≥65 | 0.45 | 0.39, 0.53 | | **<0.001** |
| **Sex (ref= male)** |  |  |  |  |
| Female | 1.53 | 1.41, 1.65 | | **<0.001** |
| **Race and ethnicity (ref= White)** | |  |  |  |
| Black | 0.87 | 0.78, 0.97 | | **0.012** |
| Hispanic | 0.87 | 0.78, 0.96 | | **0.008** |
| Other | 0.88 | 0.80, 0.97 | | **0.007** |
| **Insurance coverage (ref= Private)** | |  |  |  |
| Medicaid | 0.68 | 0.59, 0.78 | | **<0.001** |
| Medicare | 0.95 | 0.84, 1.08 | | 0.443 |
| Uninsured | 0.81 | 0.73, 0.91 | | **<0.001** |
| Other | 0.89 | 0.80, 1.00 | | **0.049** |
| **Area Deprivation index (ref= 1-2)** | |  |  |  |
| Decile: 3-8 | 1.09 | 0.96, 1.24 | | 0.175 |
| Decile: 9-10 | 1.01 | 0.87, 1.16 | | 0.919 |
| Missing | 1.07 | 0.94, 1.22 | | 0.319 |
| **Comorbidities and risk factors** | |  |  |  |
| **BMI category (ref= overweight)** | |  |  |  |
| Obesity | 1.70 | 1.55, 1.86 | | **<0.001** |
| **Coronary artery disease (ref= no)** | |  |  |  |
| Yes | 0.91 | 0.72, 1.15 | | 0.440 |
| **Heart failure (ref= no)** |  |  |  |  |
| Yes | 0.46 | 0.29, 0.72 | | **0.001** |
| **Type 2 diabetes (ref= no)** |  |  |  |  |
| Yes | 2.45 | 2.17, 2.77 | | **<0.001** |
| **Arthritis (ref= no)** |  |  |  |  |
| Yes | 0.34 | 0.18, 0.66 | | **0.002** |
| **Chronic obstructive pulmonary disease (ref= no)** | | |  |  |
| Yes | 0.80 | 0.62, 1.04 | | 0.090 |
| **Anxiety (ref= no)** |  |  |  |  |
| Yes | 1.04 | 0.88, 1.22 | | 0.663 |
| **Depression (ref= no)** |  |  |  |  |
| Yes | 1.04 | 0.86, 1.25 | | 0.698 |
| **Smoking (ref= never smoker)** | |  |  |  |
| Current smoker | 0.35 | 0.18, 0.68 | | **0.002** |
| Former smoker | 0.87 | 0.68, 1.11 | | 0.254 |
| **Charlson comorbidity index score (ref= 0)** | |  |  |  |
| 1 | 0.82 | 0.69, 0.97 | | **0.022** |
| 2+ | 1.17 | 0.93, 1.45 | | 0.174 |
| **Medication use** |  |  |  |  |
| **Antihypertensives (ref= no)** | |  |  |  |
| Yes | 1.39 | 1.20, 1.60 | | **<0.001** |
| **Statins (ref= no)** |  |  |  |  |
| Yes | 0.97 | 0.82, 1.14 | | 0.705 |
| **Metformin (ref= no)** |  |  |  |  |
| Yes | 3.45 | 2.95, 4.02 | | **<0.001** |
| **Other glucose-lowering drugs (ref= no)** |  |  |  |  |
| Yes | 2.43 | 2.05, 2.89 | | **<0.001** |
| **Antidepressants (Excludes bupropion) (ref= no)** |  |  |  |  |
| Yes | 1.39 | 1.20, 1.60 | | **<0.001** |
| **Opioids (ref= no)** |  |  |  |  |
| Yes | 0.82 | 0.74, 0.91 | | **<0.001** |
| **Provider specialty (ref= primary care)** | | | | |
| Cardiology | 1.01 | 0.84, 1.21 | | 0.944 |
| Endocrinology | 1.94 | 1.61, 2.32 | | **<0.001** |
| Nurse practitioner and physician assistant | 1.55 | 1.42, 1.69 | | **<0.001** |
| **Healthcare utilization** |  |  |  |  |
| **ER visits (ref= 0)** |  |  |  |  |
| 1+ | 0.44 | 0.40, 0.49 | | **<0.001** |
| **Hospitalization (ref= 0)** |  |  |  |  |
| 1+ | 0.55 | 0.47, 0.64 | | **<0.001** |
| **Outpatient visits (ref= 0)** |  |  |  |  |
| 1-2 | 1.41 | 1.23, 1.61 | | **<0.001** |
| 3+ | 1.30 | 1.14, 1.48 | | **<0.001** |
| Abbreviations: aOR, adjusted odds ratio, CI, confidence interval | | | |  |
| Bold values indicate significance at alpha= 0.05 | | | |  |
| *Other insurance coverage indicates those with more than one type of insurance coverage | | | |  |

**Table S5.** Factors associated with initiating newer AOMs in year 2023 (n = 2872) among a cohort of new AOM users (n= 5835) and eligible non AOM users (n= 313487).

| **Variable** | **aOR** | **95% CI** | | **p-value** |
| --- | --- | --- | --- | --- |
| **Demographics** |  |  |  |  |
| **Age (ref= 18-40)** |  |  |  |  |
| 40-65 | 1.10 | 1.01, 1.20 | | **0.028** |
| ≥65 | 0.32 | 0.27, 0.38 | | **<0.001** |
| **Sex (ref= male)** |  |  |  |  |
| Female | 1.68 | 1.55, 1.83 | | **<0.001** |
| **Race and ethnicity (ref= White)** | |  |  |  |
| Black | 0.87 | 0.78, 0.97 | | **0.016** |
| Hispanic | 0.82 | 0.74, 0.92 | | **<0.001** |
| Other | 0.91 | 0.82, 1.00 | | **0.039** |
| **Insurance coverage (ref= Private)** | |  |  |  |
| Medicaid | 0.69 | 0.60, 0.79 | | **<0.001** |
| Medicare | 0.76 | 0.65, 0.88 | | **<0.001** |
| Uninsured | 0.80 | 0.71, 0.89 | | **<0.001** |
| Other | 0.92 | 0.82, 1.03 | | **0.162** |
| **Area Deprivation index (ref= 1-2)** | |  |  |  |
| Decile: 3-8 | 0.97 | 0.86, 1.10 | | 0.618 |
| Decile: 9-10 | 0.96 | 0.84, 1.11 | | 0.604 |
| Missing | 0.95 | 0.83, 1.08 | | 0.418 |
| **Comorbidities and risk factors** | |  |  |  |
| **BMI category (ref= overweight)** | |  |  |  |
| Obesity | 1.62 | 1.48, 1.78 | | **<0.001** |
| **Coronary artery disease (ref= no)** | |  |  |  |
| Yes | 0.69 | 0.50, 0.94 | | **0.017** |
| **Heart failure (ref= no)** |  |  |  |  |
| Yes | 0.71 | 0.44, 1.16 | | 0.168 |
| **Type 2 diabetes (ref= no)** | |  |  |  |
| Yes | 1.86 | 1.61, 2.15 | | **<0.001** |
| **Arthritis (ref= no)** |  |  |  |  |
| Yes | 0.66 | 0.39, 1.10 | | 0.112 |
| **Chronic obstructive pulmonary disease (ref= no)** | | |  |  |
| Yes | 0.89 | 0.68, 1.16 | | 0.384 |
| **Anxiety (ref= no)** |  |  |  |  |
| Yes | 1.22 | 1.05, 1.42 | | **0.011** |
| **Depression (ref= no)** |  |  |  |  |
| Yes | 0.89 | 0.73, 1.08 | | 0.242 |
| **Smoking (ref= never smoker)** | |  |  |  |
| Current smoker | 0.38 | 0.21, 0.72 | | **0.003** |
| Former smoker | 0.75 | 0.57, 0.98 | | **0.038** |
| **Charlson comorbidity index score (ref= 0)** | |  |  |  |
| 1 | 0.87 | 0.71, 1.05 | | 0.144 |
| 2+ | 0.81 | 0.60, 1.10 | | 0.170 |
| **Medication use** |  |  |  |  |
| **Antihypertensives (ref= no)** | |  |  |  |
| Yes | 1.22 | 1.05, 1.42 | | **0.011** |
| **Statins (ref= no)** |  |  |  |  |
| Yes | 0.91 | 0.75, 1.09 | | 0.308 |
| **Metformin (ref= no)** |  |  |  |  |
| Yes | 2.34 | 1.94, 2.83 | | **<0.001** |
| **Other glucose-lowering drugs (ref= no)** |  |  |  |  |
| Yes | 1.75 | 1.39, 2.21 | | **<0.001** |
| **Antidepressants (Excludes bupropion) (ref= no)** |  |  |  |  |
| Yes | 0.89 | 0.73, 1.08 | | 0.242 |
| **Opioids (ref= no)** |  |  |  |  |
| Yes | 0.84 | 0.76, 0.93 | | **0.001** |
| **Provider specialty (ref= primary care)** | | | | |
| Cardiology | 0.99 | 0.81, 1.20 | | 0.903 |
| Endocrinology | 1.87 | 1.54, 2.29 | | **<0.001** |
| Nurse practitioner and physician assistant | 1.65 | 1.51, 1.80 | | **<0.001** |
| **Healthcare utilization** |  |  |  |  |
| **ER visits (ref= 0)** |  |  |  |  |
| 1+ | 0.51 | 0.47, 0.57 | | **<0.001** |
| **Hospitalization (ref= 0)** |  |  |  |  |
| 1+ | 0.54 | 0.46, 0.63 | | **<0.001** |
| **Outpatient visits (ref= 0)** |  |  |  |  |
| 1-2 | 1.24 | 1.09, 1.41 | | **0.001** |
| 3+ | 1.23 | 1.09, 1.40 | | **0.001** |
| Abbreviations: aOR, adjusted odds ratio, CI, confidence interval | | | |  |
| Bold values indicate significance at alpha= 0.05 | |  |  |  |
| *Other insurance coverage indicates those with more than one type of insurance coverage | | | | |

**Table S6.** Factors associated with initiating tirzepatide (n= 712) among a cohort patients without type 2 diabetes, receiving semaglutide (n= 1765) or tirzepatide (n= 712) in 2023.

| **Variable** | **aOR** | **95% CI** | | **p-value** |
| --- | --- | --- | --- | --- |
| **Demographics** |  |  |  |  |
| **Age (ref= 18-40)** |  |  |  |  |
| 40-65 | 1.00 | 0.82, 1.21 | | 0.972 |
| ≥65 | 1.18 | 0.78, 1.78 | | 0.436 |
| **Sex (ref= male)** |  |  |  |  |
| Female | 1.13 | 0.92, 1.40 | | 0.246 |
| **Race and ethnicity (ref= White)** | |  |  |  |
| Black | 0.65 | 0.49, 0.87 | | 0.003 |
| Hispanic | 0.75 | 0.57, 0.98 | | 0.035 |
| Other | 1.03 | 0.83, 1.28 | | 0.791 |
| **Insurance coverage (ref= Private)** | |  |  |  |
| Medicaid | 0.85 | 0.61, 1.19 | | 0.347 |
| Medicare | 0.80 | 0.53, 1.22 | | 0.305 |
| Uninsured | 1.01 | 0.76, 1.33 | | 0.965 |
| Other | 0.95 | 0.72, 1.25 | | 0.73 |
| **Area Deprivation index (ref= 1-2)** | |  |  |  |
| Decile: 3-8 | 1.05 | 0.78, 1.41 | | 0.746 |
| Decile: 9-10 | 0.96 | 0.69, 1.34 | | 0.822 |
| Missing | 0.99 | 0.72, 1.36 | | 0.961 |
| **Comorbidities and risk factors** | |  |  |  |
| **BMI category (ref= overweight)** | |  |  |  |
| Obesity | 1.12 | 0.88, 1.43 | | 0.338 |
| **Coronary artery disease (ref= no)** | |  |  |  |
| Yes | 2.34 | 0.83, 6.56 | | 0.107 |
| **Type 2 diabetes (ref= no)** | |  |  |  |
| Yes |  |  |  |  |
| **Arthritis (ref= no)** |  |  |  |  |
| Yes | 1.27 | 0.87, 1.85 | | 0.208 |
| **Chronic obstructive pulmonary disease (ref= no)** | | |  |  |
| Yes | 0.99 | 0.48, 2.03 | | 0.979 |
| **Anxiety (ref= no)** |  |  |  |  |
| Yes | 1.27 | 0.87, 1.85 | | 0.208 |
| **Depression (ref= no)** |  |  |  |  |
| Yes | 0.60 | 0.35, 1.02 | | 0.061 |
| **Charlson comorbidity index score (ref= 0)** | |  |  |  |
| 1 | 0.74 | 0.37, 1.48 | | 0.392 |
| 2+ | 2.44 | 0.83, 7.19 | | 0.106 |
| **Medication use** |  |  |  |  |
| **Antihypertensives (ref= no)** | |  |  |  |
| Yes | 1.25 | 0.88, 1.79 | | 0.211 |
| **Statins (ref= no)** |  |  |  |  |
| Yes | 0.54 | 0.33, 0.89 | | **0.017** |
| **Metformin (ref= no)** |  |  |  |  |
| Yes | 1.04 | 0.65, 1.68 | | 0.864 |
| **Antidepressants (Excludes bupropion) (ref= no)** |  |  |  |  |
| Yes | 1.25 | 0.88, 1.79 | | 0.211 |
| **Opioids (ref= no)** |  |  |  |  |
| Yes | 0.98 | 0.74, 1.28 | | 0.856 |
| **Provider specialty (ref= primary care)** | | | | |
| Cardiology | 1.13 | 0.68, 1.88 | | 0.638 |
| Endocrinology | 1.12 | 0.68, 1.84 | | 0.651 |
| Nurse practitioner and physician assistant | 1.13 | 0.92, 1.38 | | 0.261 |
| **Healthcare utilization** |  |  |  |  |
| **ER visits (ref= 0)** |  |  |  |  |
| 1+ | 0.76 | 0.60, 0.96 | | **0.021** |
| **Hospitalization (ref= 0)** |  |  |  |  |
| 1+ | 1.04 | 0.68, 1.59 | | 0.857 |
| **Outpatient visits (ref= 0)** |  |  |  |  |
| 1-2 | 0.68 | 0.51, 0.92 | | **0.013** |
| 3+ | 0.76 | 0.57, 1.01 | | 0.062 |
| Abbreviations: aOR, adjusted odds ratio, CI, confidence interval  Bold values indicate significance at alpha= 0.05  *Other insurance coverage indicates those with more than one type of insurance coverage | | | | |
